# Evaluation of a real time machine learning sepsis risk algorithm for Emergency Department waiting rooms (SAFE-WAIT)

**DOI:** 10.1101/2025.06.23.25330154

**Authors:** Gladis Kabil, Steven A Frost, Audrey P Wang, Minh Trang Hoang, Pramod Chandru, Michelle Moscova, Amith Shetty

## Abstract

**Objectives:** To evaluate and compare the real-time Sepsis risk Artificial intelligence algorithm For Emergency department WAITing room (SAFE-WAIT) model with the standard SIRS-based sepsis alert in recognizing and managing sepsis.

**Study Design:** A retrospective analysis of the AI algorithm predicting sepsis risk using an ongoing emergency department sepsis archive.

**Setting and Participants:** Adults presenting to a metropolitan emergency department in Western Sydney between July 2022 and June 2024 who received either a SAFE-WAIT risk category and/or the standard sepsis alert.

**Main Outcome Measures:** The primary outcomes: recognition of the development of sepsis and septic shock. Secondary outcomes: the time to physician review and initial antibiotic administration.

**Results:** Among 108,401 patients analysed, 104,904 received a SAFE-WAIT risk category. Of these, 5,149 (4.9%) had a confirmed sepsis diagnosis, and 312 (3%) developed septic shock. SAFE-WAIT categorized 270 (86.5%) of septic shock patients as high risk at triage, with event onset at 92 minutes (Inter Quartile Range: 17.25–233.00). Adjusted β-coefficients showed significantly faster antibiotic administration in moderate and high-risk SAFE-WAIT groups (Moderate: –58.81 minutes, 95% Confidence Interval (CI): –72.26 to –35.80, p < ; High: –116.99 minutes, 95% CI: –135.34 to –98.65, p < 0.001). High-risk patients had a slightly shorter time to first physician review (β = –3.5 minutes, 95% CI: –5.41 to –1.58, p < 0.001). These effects of SAFE WAIT grouping on the time to antibiotics and clinician review were both modified by triage category (*p* for interaction < 0.001).

**Conclusions:** SAFE-WAIT effectively predicts sepsis-related adverse events at triage, positively impacting sepsis management. These findings underscore the potential role of AI-augmented clinical practice.

## Introduction

Sepsis recognition remains a significant challenge, with increasing emergency department (ED) wait times contributing to delays in identification and treatment.^(1)^ Sepsis patients often deteriorate rapidly, requiring swift diagnosis and intervention to improve survival. Artificial intelligence (AI) tools have demonstrated effectiveness in predicting patient deterioration in general wards, often outperforming conventional methods.^(2)^ However, the performance of sepsis screening tools, including AI-based algorithms, in the ED has been inconsistent. A recent study^(3)^ comparing physician gestalt to existing sepsis screening scores—including Systemic Inflammatory Response Syndrome (SIRS), Sequential Organ Failure Assessment (SOFA), quick SOFA (qSOFA), Modified Early Warning Score (MEWS), and a machine learning (ML) model—found that physician gestalt within the first 15 minutes of patient assessment outperformed other screening methods in identifying sepsis. This underscores the need for enhanced sepsis screening tools capable of early risk identification, facilitating timely physician review and antibiotic administration.(^2,3^) This study evaluates the performance of the real-time Sepsis risk AI screening algorithm for Emergency Department WAITing room (SAFE-WAIT) in sepsis recognition and management. We compared SAFE-WAIT’s performance with the existing SIRS-based ED electronic medical record (EMR) alert (WSLHD sepsis alert), hypothesizing that SAFE-WAIT enables earlier risk categorization and treatment initiation.

Primary outcomes include recognition of sepsis and septic shock development, with secondary measures assessing time to first physician review and antibiotic administration.

## Methods

### Study design and setting

This study was a retrospective analysis of a large data set of ED patient encounters (n=453,121) presenting to metropolitan hospital in Western Sydney (SMEDSA)^(4)^. Data from patients who presented to the ED during the prospective SAFE-WAIT clinical deployment phase between July 2022 to June 2024 were collected as part of the ongoing sepsis longitudinal study. Ethics approval was obtained from the WSLHD Ethics Committee (Reference numbers 2019/ETH09607 and 2020/ETH03269). The program was evaluated and managed in keeping with the principles of the NSW Artificial Intelligence Assurance Approach and adhered to the NSW AI assessment framework.

### SAFE WAIT Model development

The SAFE-WAIT is an Extreme Gradient Boosting (XG-Boost) ML model^(5)^ developed to predict the probability of the risk of developing sepsis for patients in the ED waiting room. Data from January 2017 to December 2019 collected from four Western Sydney metropolitan EDs as part of the SMEDSA initiative were used for model training to provide a comprehensive historical context.^6,7^ A range of clinical data—including patient demographics, triage assessments, medical history, laboratory results, interventions, and outcomes was extracted from the electronic Medical Records (eMR). This information was used to code the patient cohort with a confirmed sepsis diagnosis, based on the Sepsis-3 criteria^(6)^, and for model training.

### Model deployment

Model development was followed by a silent trial which was conducted between January and June 2022 at one of four designated test hospitals. As the model was designed to risk-stratify sepsis in the waiting room, only features readily available at triage such as vital signs, age, and gender, were used as input variables. Technical details of the model development and evaluation of the performance of the SAFE-WAIT model during the silent trial have been described in prior publications.^6,7^ At the study site, the SAFE-WAIT algorithm was implemented using educational and training sessions conducted by the clinical model development team and multidisciplinary group of emergency clinicians. The clinical workflow of the implementation site is depicted in **Figure 1**. The SAFE-WAIT model identifies the patients at-risk of developing sepsis in the waiting room, once the triage nurse inputs the vital signs in the eMR triage notes. SAFE-WAIT model categorized patients into three groups – low-risk, medium-risk and high-risk of developing sepsis. SAFE-WAIT categories were then displayed on the dashboard parallel to the eMR available to all clinicians (**Figure 2**). Since the screening focused mainly on the waiting room, the triage and waiting room support nurse were primarily trained to review the waiting room risk dashboard regularly with advice to monitor high-risk patients closely (repeated vitals measurement), initiate earlier investigations e.g. lactate measurements or seek early physician review when necessary.

**Figure 1:**
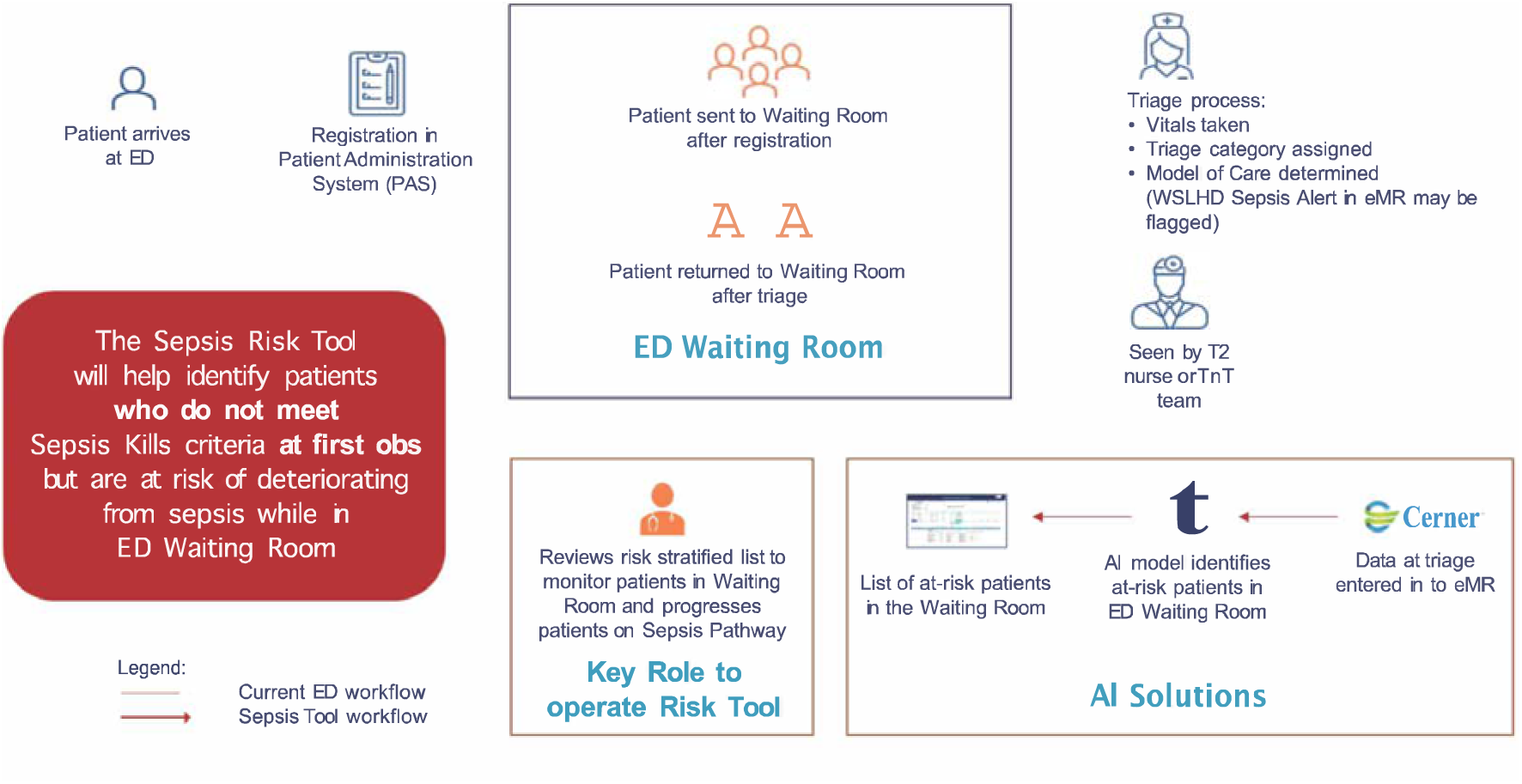
Clinical Workflow of implementation site

**Figure 2:**
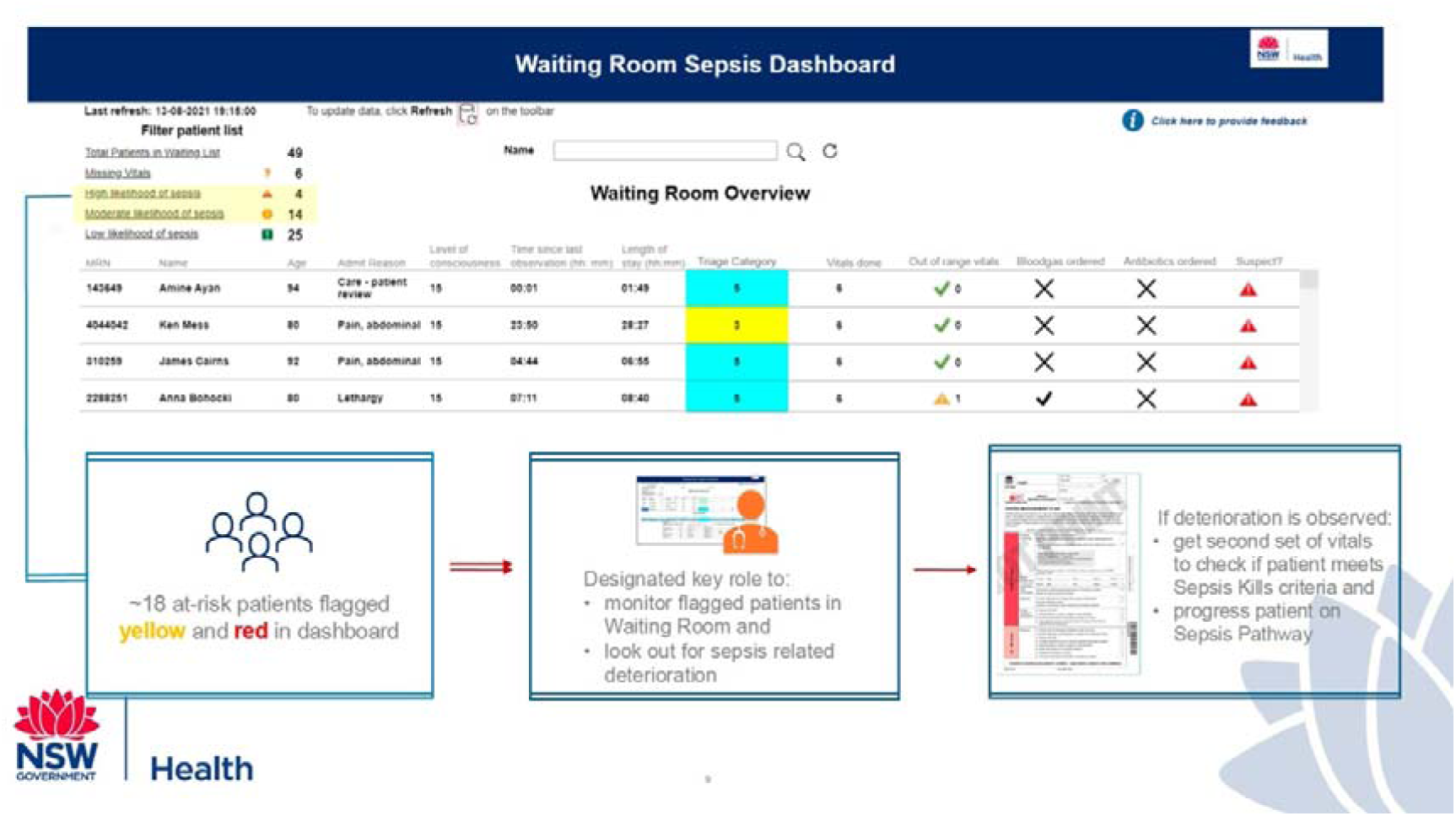
SAFE-WAIT dashboard display

### Participant selection

In our analysis, we included patients older than 16 years presenting to the metropolitan hospital where the SAFE-WAIT algorithm was deployed. Patients who received a triage category 1 were excluded, as they typically bypass the standard triage process to resuscitation bays in ED. Additionally, we excluded those who did not have a complete set of vital signs observations recorded at triage (**Figure 3**) as SAFE-WAIT risk category could not be calculated for those patients. Thus, we included patients who received the SAFE-WAIT risk category in the waiting room and/or those who received the previously implemented ED electronic medical record (EMR) SIRS-based sepsis alert (WSLHD sepsis alert) ^(8)^.

**Figure 3:**
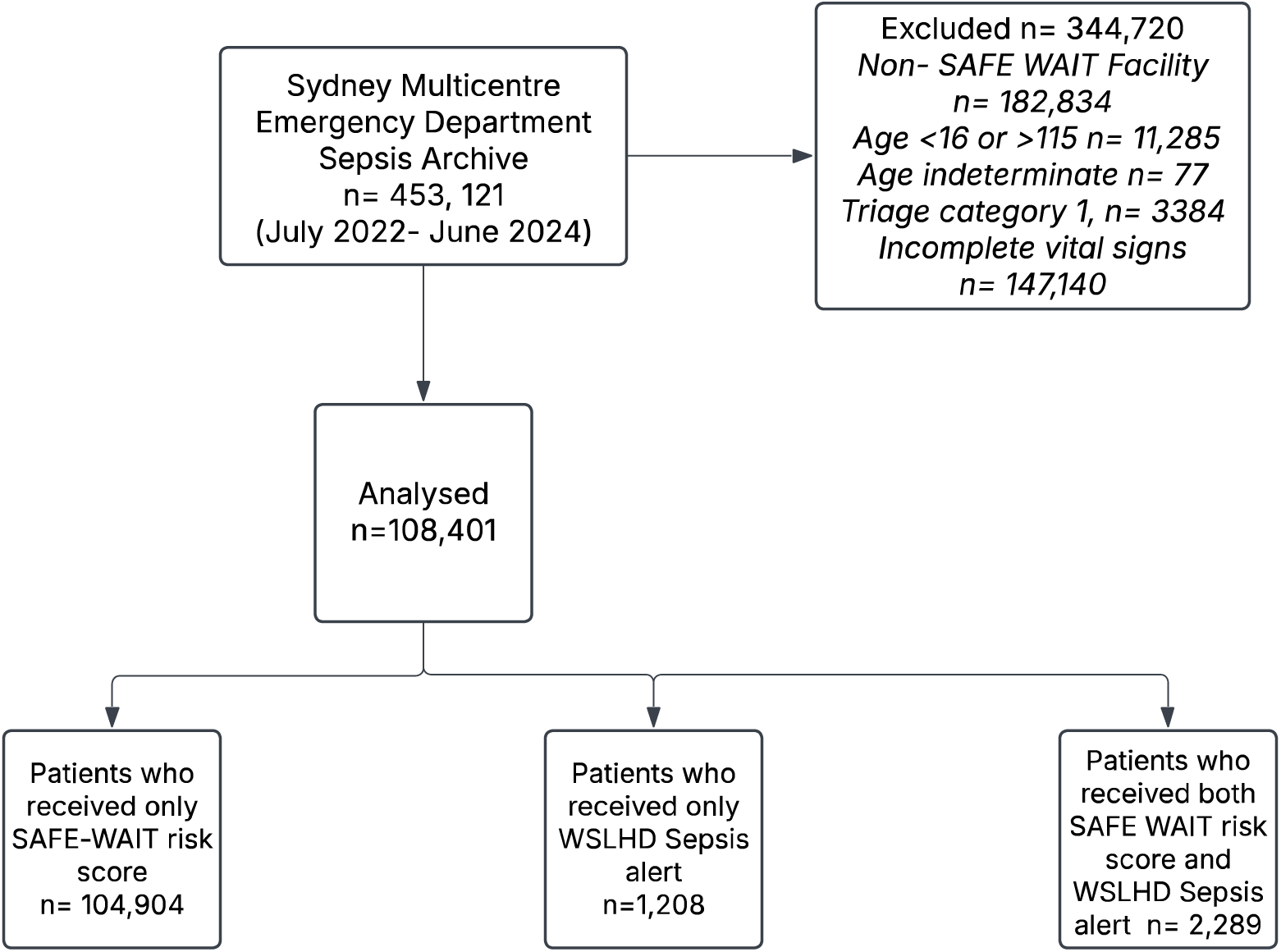
Flowchart of patient encounters included

### Statistical analysis

Data management and analysis were performed using the R language for statistical analysis software (version 4.0.3, R Foundation for Statistical Computing, Vienna, Austria). Descriptive summaries and outcomes were reported for patients who received a SAFE-WAIT category and were stratified as (a) low risk (b) moderate risk and (c) high risk, and those who received a WSLHD Sepsis alert only. Multivariable linear regression models were used to identify the association between SAFE-WAIT risk category and the time to antibiotics, and time to the first physician review. Adjusted estimate of effects is presented as β-co-efficient, 95% Confidence Intervals (CIs). To identify potential confounders, all covariates were included in the models initially, and a stepwise approach (forward and backward elimination) was used to develop the final models. In patients with confirmed sepsis in ED, we measured the association between the assigned SAFE-WAIT category and the WSLHD sepsis alert, with the time to physician review, time to first dose of antibiotics, ICU admission and all-cause in-hospital mortality rates. The adjusted effects estimates for ICU admission and all-cause in-hospital mortality from the logistic regression were reported as Odds Ratio and 95% CIs. Triage category was explored as a potential effect modifier, with a p-value < 0.10 set as significant; therefore, results have been presented for each triage category separately, with a *p*-value for an interaction term. The effect of triage categories was examined in both the linear and logistic regression models.

To evaluate SAFE-WAIT, we performed decision curve analysis to calculate a clinical “net benefit” for SAFE-WAIT in comparison to default strategies of treating all or no patients.^(8)^ The “net benefit” value informed the benefit of correctly predicting the risk of developing sepsis (true positive) and the relative harm of incorrectly predicting risk of developing sepsis (false positive) – over a range from threshold probability of the outcome.^(9)^ The decision curve was plotted for all patients receiving SAFE-WAIT alert and for patients in each risk category of SAFE-WAIT to identify the group with highest net benefit. To compare SAFE-WAIT performance with the existing WLSHD Sepsis alert, we reported the standard performance metrics.

## Results

A total of 108,401 patient encounters were analysed, excluding cases from three hospitals without SAFE-WAIT implementation, as well as those with incomplete data on age (n=77) and vital signs (n=147,140). Additionally, 3,384 patients classified as Category 1 were excluded. Among the included encounters, 104,904 received only a SAFE-WAIT risk category, 1,208 triggered the WSLHD Sepsis alert, and 2,289 were flagged by both systems (**Figure 3**).

Older patients were more frequently classified as high-risk by SAFE-WAIT (38% of patients ≥ 65 years old) and received the WSLHD Sepsis alert (43% of patients ≥ 65 years old) (**Table 1**). Median triage times were shorter for high-risk patients (6.0 minutes, Inter Quartile Range (IQR): 3.0–12.0) compared to low-risk patients (9.0 minutes, IQR: 4.0–17.0). Most high-risk patients (94%) received a triage category of 2–3.

**Table 1.** Characteristics of patients based on SAFE WAIT risk and WSLHD Sepsis Alert.

|  | **Low Risk**<br>(N=59617) | **Moderate Risk**<br>(N=35561) | **High Risk**<br>(N=9726) | **WSLHD Sepsis Alert**<br>(N=1208) |
| --- | --- | --- | --- | --- |
| **Male, N (%)** | 26320 (44%) | 19326 (54%) | 5638 (58%) | 616 (51%) |
| **Age Category, N (%)** |  |  |  |  |
| Y16-Y35 | 19974 (34%) | 13497 (38%) | 2645 (27%) | 249 (21%) |
| Y36-Y65 | 27569 (46%) | 13696 (38%) | 3429 (35%) | 443 (37%) |
| Y66-Y115 | 12074 (20%) | 8368 (24%) | 3652 (38%) | 516 (43%) |
| **Time to triage, in minutes Median (IQR)** | 9.0 (4.0 to 17.0) | 7.0 (3.0 to 14.0) | 6.0 (3.0 to 12.0) | 8.0 (3.0 to 15.0) |
| **Triage Category, N (%)** |  |  |  |  |
| 2-3 | 33978 (57%) | 29617 (83%) | 9117 (94%) | 1198 (99%) |
| 4-5 | 25639 (43%) | 5944 (17%) | 609 (6%) | 10 (1%) |
| **Presentation hour, N (%)** |  |  |  |  |
| 0700-1500 h | 29617 (50%) | 16500 (46%) | 4387 (46%) | 519 (43%) |
| 1500-2300 h | 22784 (38%) | 14078 (40%) | 3930 (40%) | 553 (46%) |
| 2300-0700 h | 7216 (12%) | 4983 (14%) | 1409 (14%) | 136 (11%) |

Among SAFE-WAIT assessed patients, 24% (2334) of high-risk cases and 47% (569) of WSLHD Sepsis alert cases developed confirmed sepsis (**Table 2**). Septic shock incidence rose with risk category: <1% in low/moderate SAFE-WAIT groups, 3% in high-risk SAFE-WAIT patients, and 8% in WSLHD alert patients. Median time to septic shock onset (Systolic Blood Pressure ≤ 90 mmHg) was 92 minutes (IQR: 17.25–233.00) for SAFE-WAIT high-risk patients.

**Table 2:** Confirmed Sepsis, WSLHD Sepsis Alert and SAFE-WAIT category.

|  | SAFE-WAIT<br>N= 104,909 |  |  | WSLHD Sepsis Alert |
| --- | --- | --- | --- | --- |
|  | Low Risk<br>N= 59617 | Moderate Risk<br>N=35561 | High Risk<br>N=9726 | N=1208 |
| **Confirmed Sepsis,**<br>**N (%)** | 799 (1%) | 2016 (6%) | 2334 (24%) | 569 (47%) |
| **Septic shock**<br>**N (%)** | 3 (<1%) | 39 (<1%) | 270 (3%) | 95 (8%) |
| **Time to first**<br>**physician review**<br>**min, Median**<br>**(IQR)** | 94 (44 to 182) | 105 (48 to 202) | 95 (44 to 189) | 75 (40 to 140) |
| **Time to Systolic**<br>**BP <90 (min),**<br>**Median (IQR)** | 249 (92.75 to<br>424.25) | 153 (34 to 370) | 92 (17.25 to<br>233.00) | 142 (44.25 to<br>293.50) |
| **Time to**<br>**antibiotics hrs,**<br>**Median (IQR)** | 3.7 (2.0 to 6.8)<br>(n= 6972[11.7%]) | 5.1 (2.7 to 9.1)<br>(n= 6442[18.1%]) | 4.5 (2.4 to 7.8)<br>(n=4164[42.8%]) | 2.8 (1.5 to 5.2)<br>(n=1026[84.9%]) |
| **ICU admission,**<br>**N (%)** | 159 (<1%) | 289 (1%) | 455 (5%) | 123 (10%) |
| **All-Cause In-**<br>**hospital**<br>**mortality,**<br>**N (%)** | 131 (<1%) | 376 (1%) | 457 (5%) | 84 (7%) |
| Number of valid records for Time to antibiotics, N =17578; n = number of patients who received antibiotic within the risk group; Time to first physician review, N=70841 |  |  |  |  |

High-risk SAFE-WAIT patients had slightly earlier physician review (β = –3.5 minutes, 95% Confidence Intervals (CI): –5.41 to –1.58; p < 0.001). Moderate-risk patients in triage categories 4–5 experienced more significant delays (β = 10.62 minutes, 95% CI: 6.71–14.53; p < 0.001). A statistically significant interaction (p < 0.001) suggested triage category influenced physician response time.

After adjusting for age, gender, presentation hour, and SOFA scores (**Table 3**), antibiotic administration occurred significantly faster in moderate and high-risk SAFE-WAIT groups compared to low-risk patients. Antibiotics were delivered earlier in the moderate-risk group by 58.81 minutes (95% CI: –72.26 to –35.80; p < 0.001) and in the high-risk group by 116.99 minutes (95% CI: –135.34 to –98.65; p < 0.001). When stratified by triage category, high-risk patients in triage category 2–3 received antibiotics 83.60 minutes earlier (95% CI: – 108.84 to –58.37; p < 0.001), while those in triage 4–5 received them 186.37 minutes later (95% CI: 115.86 to 256.88; p < 0.001). A significant interaction (p < 0.001) between SAFE-WAIT risk category and triage level indicated that risk level affected antibiotic timing differently across triage categories. Similar trends were observed when analysis was restricted to patients with confirmed sepsis in the ED (**Appendix 1**).

**Table 3.** Association between SAFE WAIT category and time to antibiotics, and first physician review.

|  | SAFE WAIT Category |  |  | TRIAGE Category |
| --- | --- | --- | --- | --- |
| Variable | Low Risk | Moderate Risk | High Risk | 2-3 Vs 4-5 (Ref:4-5) |
| **Time to antibiotics in hrs**<br>(Adjusted $\beta$ Co-efficient, 95% CI),<br>P value | | | | |
| All patients | Ref | 54.03 (35.80 to 72.26),<br><0.001 | -58.81 (35.80 to -72.26), <0.001 | -116.99 (-135.34 to -98.65), <0.001 |
| Triage Category 2-3 | Ref | 40.45 (17.80 to 63.11),<br><0.001 | -83.60 (-108.84 to -58.37), <0.001 |  |
| Triage Category 4-5 | Ref | 39.57 (8.59 to 70.56),<br>0.0123 | 186.37 (115.86 to 256.88), <0.001 |  |
| P value for interaction | < 0.001 |  |  |  |
| **Time to first physician review in min**<br>(Adjusted $\beta$ Co-efficient, 95% CI) | | | | |
| All patients | Ref | 8.46 (6.55 to 10.37),<br>< 0.001 | -0.91 (-4.08 to 2.25),<br>0.573 | -3.5 (-5.41 to -1.58),<br>< 0.001 |
| Triage category 2-3 | Ref | 6.29 (4.09 to 8.49),<br>< 0.001 | -2.23 (-5.62 to 1.14),<br>0.194 |  |
| Triage Category 4-5 | Ref | 10.62 (6.71 to 14.53),<br>< 0.001 | 18.6 (7.96 to 29.24),<br>< 0.001 |  |
| P value for interaction | < 0.001 |  |  |  |
| All variables are adjusted for age, gender, presentation hour and SOFA scores |  |  |  |  |

ICU admission and all-cause in-hospital mortality followed similar trends, with increased odds in high-risk patients. Adjusted odds ratio (OR) for high-risk group patients for ICU admission was 1.77 (95% CI: 1.28 to 2.45; p < 0.001), while adjusted OR for in-hospital mortality was 2.26 (95% CI: 1.52 to 3.34; p < 0.001).

On average, for every 4.8 patients flagged high-risk, an additional patient with an adverse outcome—such as sepsis or septic shock—was identified.

**Table 4** describes the performance metrics of SAFE-WAIT in comparison with the existing WSLHD Sepsis alert. The SAFE-WAIT model demonstrated significantly higher sensitivity (0.86, 95% CI: 0.85–0.87) compared to the WSLHD alert (0.13, 95% CI: 0.13–0.14), indicating a greater ability to identify patients at risk of developing sepsis. While SAFE-WAIT exhibited a higher false positive rate (0.42), it also had a markedly lower false negative rate (0.14) compared to the WSLHD alert (0.02 and 0.87, respectively). These findings suggest that SAFE-WAIT is better suited for capturing at-risk patients in the emergency department waiting room, enabling earlier clinical review and closer monitoring. The decision curve analysis (**Figure 4**) further supports the utility of SAFE-WAIT in the ED waiting room. Across a wide range of threshold probabilities, the model demonstrated greater net benefit than both default strategies of treating all patients or none. Notably, SAFE-WAIT provided the highest clinical utility at lower thresholds, reflecting its ability to predict the risk of developing sepsis early while maintaining reasonable false positive rates.

**Figure 4:**
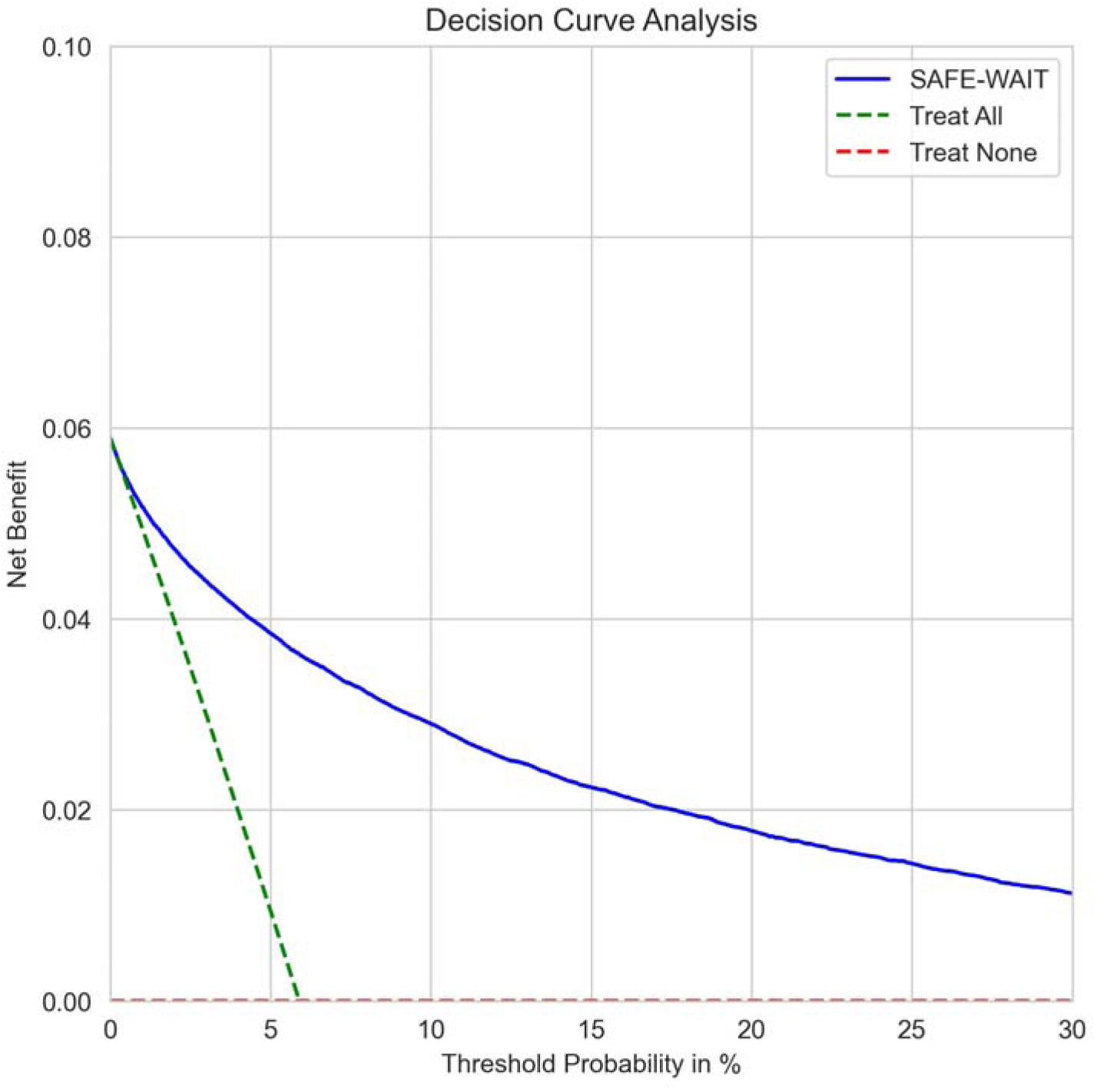
Decision curve analysis for the performance of SAFE WAIT in identifying patient at risk of sepsis

**Figure 5:**
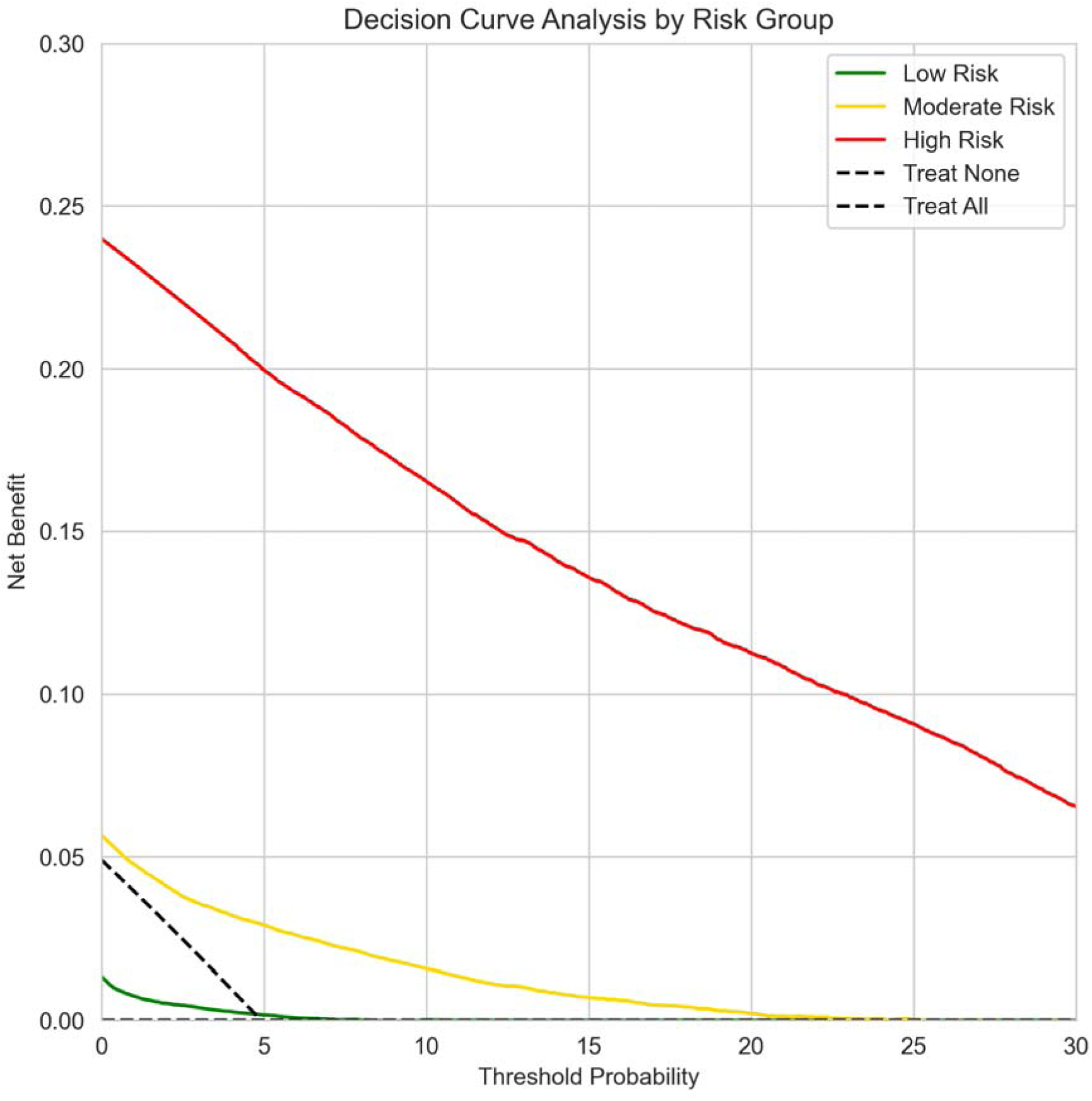
Decision curve analysis for the performance of SAFE WAIT in identifying patient at risk of sepsis stratified by each risk group

**Table 4.** Performance metrics of SAFE-WAIT vs. WSLHD Sepsis alert in all patients.

| Model | Sensitivity<br>(95% CI) | Specificity<br>(95% CI) | Positive<br>predicting<br>value<br>(95% CI) | Negative<br>predicting<br>value<br>(95% CI) | Accuracy | False<br>Positive<br>Rate | False<br>Negative<br>Rate |
| --- | --- | --- | --- | --- | --- | --- | --- |
| SAFE-WAIT | 0.86 (0.85 – 0.87) | 0.58 (0.58, 0.58) | 0.11 (0.1, 0.11) | 0.99 (0.99, 0.99) | 0.60 | 0.42 | 0.14 |
| WSLHD sepsis alert | 0.13 (0.13, 0.14) | 0.98 (0.98, 0.98) | 0.38 (0.37, 0.4) | 0.94 (0.94, 0.94) | 0.93 | 0.02 | 0.87 |

To further explore the clinical utility of the model, we examined net benefit within three risk categories (low, moderate, and high) based on the model’s own predicted probabilities (**Figure 6**). The highest net benefit was observed in the high-risk group, across all thresholds. The moderate-risk group demonstrated modest benefit, particularly at lower thresholds, while the low-risk group showed minimal net benefit.

## Discussion

Our results suggest that the SAFE-WAIT AI sepsis risk screening tool effectively stratifies patients at risk of sepsis at emergency department (ED) triage. The trial demonstrated significant improvements in time to antibiotics and time to first physician review across SAFE-WAIT risk categories and triage levels.

In this Australian-first implementation of a real-time sepsis risk AI tool, SAFE-WAIT was deployed to assist waiting room and triage nurses in monitoring ED patients. The SAFE-WAIT dashboard enabled nurses to identify patients requiring closer monitoring, which could result in instances of more frequent vital signs checks, lactate measurements, clinical investigations, or early physician reviews upon signs of deterioration. While AI predictive tools have proven effective in Intensive Care Units (ICUs) ^(10)^ and general wards^(2)^—showing accurate sepsis predictions and improved intervention times—studies in ED settings have shown variable results with time to intervention. (^11,12^)

Unlike AI models in ICUs and general wards, which utilize serial vital signs, pathology results, and disease severity scores (SOFA), this study implemented SAFE-WAIT using only triage-collected data, such as vital signs and demographics. This approach enhances generalizability across different ED settings.

Early recognition of sepsis, monitoring for deterioration, and timely interventions—including prompt antibiotic administration—are critical in improving patient outcomes.(^13,14,15^) However, challenges such as overcrowding, staffing shortages, and complex presentations in EDs hinder early recognition and intervention.(^16,17,18^) As extended ED wait times contribute to delays, recognising high-risk patients before physician review becomes imperative. SAFE-WAIT provides real-time risk updates with each vital sign measurement, offering nurses a trend analysis of patient deterioration.

Our study site previously employed a SIRS-based sepsis alert, which flagged patients meeting predefined criteria, placing them on the Clinical Excellence Commission (CEC) Sepsis pathway. Interestingly, not all patients who developed sepsis in ED were flagged by this alert. However, patients who triggered the WSLHD sepsis alert had higher rates of septic shock, ICU admission, and in-hospital mortality, suggesting that patients meeting these criteria may be sicker or experience altered disease trajectories. Utilizing these alerts to trigger sepsis pathway interventions remains prudent and aligns with findings from other real-time predictor models recently implemented in EDs.^(19)^ Electronic Medical Record (EMR)-based rules engines, which typically rely on binomial cut-offs or threshold-based logic (e.g., vital sign values above or below predefined limits), are inherently limited in their ability to capture the nuanced, nonlinear interactions that underlie patient deterioration. These deterministic systems often lack sensitivity and specificity, particularly in complex clinical scenarios where multiple variables interact in non-additive ways. In contrast, artificial intelligence (AI) and machine learning (ML) algorithms are capable of modelling high-dimensional data and uncovering complex polynomial and temporal interactions among physiological parameters, laboratory results, and clinical histories. This allows for more accurate and dynamic risk stratification of deteriorating patients, enabling earlier and more targeted interventions. As such, AI/ML models represent a significant advancement over traditional rules-based systems in predictive clinical analytics.^(20)^

The stratified decision curve analysis indicates that SAFE-WAIT provides the greatest net clinical benefit in patients classified as high risk. These findings support that patients of high-risk should be closely monitored for timely intervention while low-risk patients may not benefit meaningfully from additional interventions.

In our study, most septic shock patients were categorized as high-risk by SAFE-WAIT at triage. Notably, this risk stratification occurred within six minutes after presentation, approximately 90 minutes earlier than the onset of clinical signs of septic shock. This aligns with findings from Kim et al. ^(21)^ where a machine learning algorithm at triage outperformed traditional sepsis screening tools. Our analysis suggests that one in five high-risk patients would benefit from close monitoring and early interventions in the waiting room (Number needed to treat = 4.8).

SAFE-WAIT differs from traditional electronic medical alerts used in sepsis care. Prior studies(^22,23,24^) focused on identifying sepsis through clinical criteria, followed by electronic alerts triggering escalation over several hours. SAFE-WAIT, however, aimed to predict sepsis risk before the first physician review. Of particular importance are our findings of reduced physician review time and faster antibiotic administration in patients categorized as high-risk by SAFE-WAIT. However, our results also indicate that triage category significantly influences physician review times and antibiotic administration, highlighting the crucial role of triage nurses in clinical assessment. Enhancing the AI algorithm with triage assessment notes will be the focus of future research.

Future research should explore AI-enhanced triage algorithms to further optimize risk stratification and triaging processes. While SAFE-WAIT focused on sepsis risk in waiting room patients, this approach could be extended to broader patient deterioration screening for those awaiting care.

## Limitations

This study has several limitations that should be considered when interpreting the findings. Firstly, the retrospective observational design limits causal inference, and unmeasured confounders such as clinician experience, patient complexity, and departmental workload that may have influenced outcomes. Secondly, the SAFE-WAIT algorithm’s effectiveness was contingent on the completeness of triage data, particularly vital signs. Patients without a full set of observations were excluded, potentially introducing selection bias. Although data completeness improved over time with increased awareness, this may have introduced temporal bias.

Thirdly, the study could not account for ED crowding metrics or staffing levels, both of which are known to impact patient flow and timely care delivery. Importantly, the study relied on eMR timestamps to measure clinical events such as time to physician review and antibiotic administration. These timestamps may not accurately reflect real-time clinical processes. For instance, antibiotics may be prescribed by physicians without formally assigning themselves to the patient in the eMR, leading to potential underestimation of physician involvement or delays in recorded intervention times. This discrepancy may affect the interpretation of time-sensitive outcomes.

Additionally, the uptake of the SAFE-WAIT tool was lower than anticipated mainly due to the parallel presentation of dashboard rather than integration into the eMR. This limited adoption may have attenuated the potential impact of the intervention and reflects the need for further investigation into barriers to clinical integration.

Lastly, the generalisability and scalability of findings may be limited to similar urban ED settings with comparable digital infrastructure and triage workflows. Future studies should explore prospective validation, broader implementation strategies, and integration with clinician assessments to enhance the utility and adoption of AI-based risk stratification tools.

## Conclusions

This study demonstrates that the SAFE-WAIT AI algorithm can effectively identify patients at risk of sepsis at the point of triage, enabling earlier clinical intervention and improved time to antibiotics. By leveraging routinely collected triage data, SAFE-WAIT supports timely decision-making in the emergency department, particularly in high-demand settings. These findings underscore the potential of AI-augmented clinical practice to enhance early recognition and management of critical illness, reinforcing the value of integrating predictive tools into frontline care pathways.

## Data Availability

Data used in the manuscript are unavailable to public due to ethical restrictions.

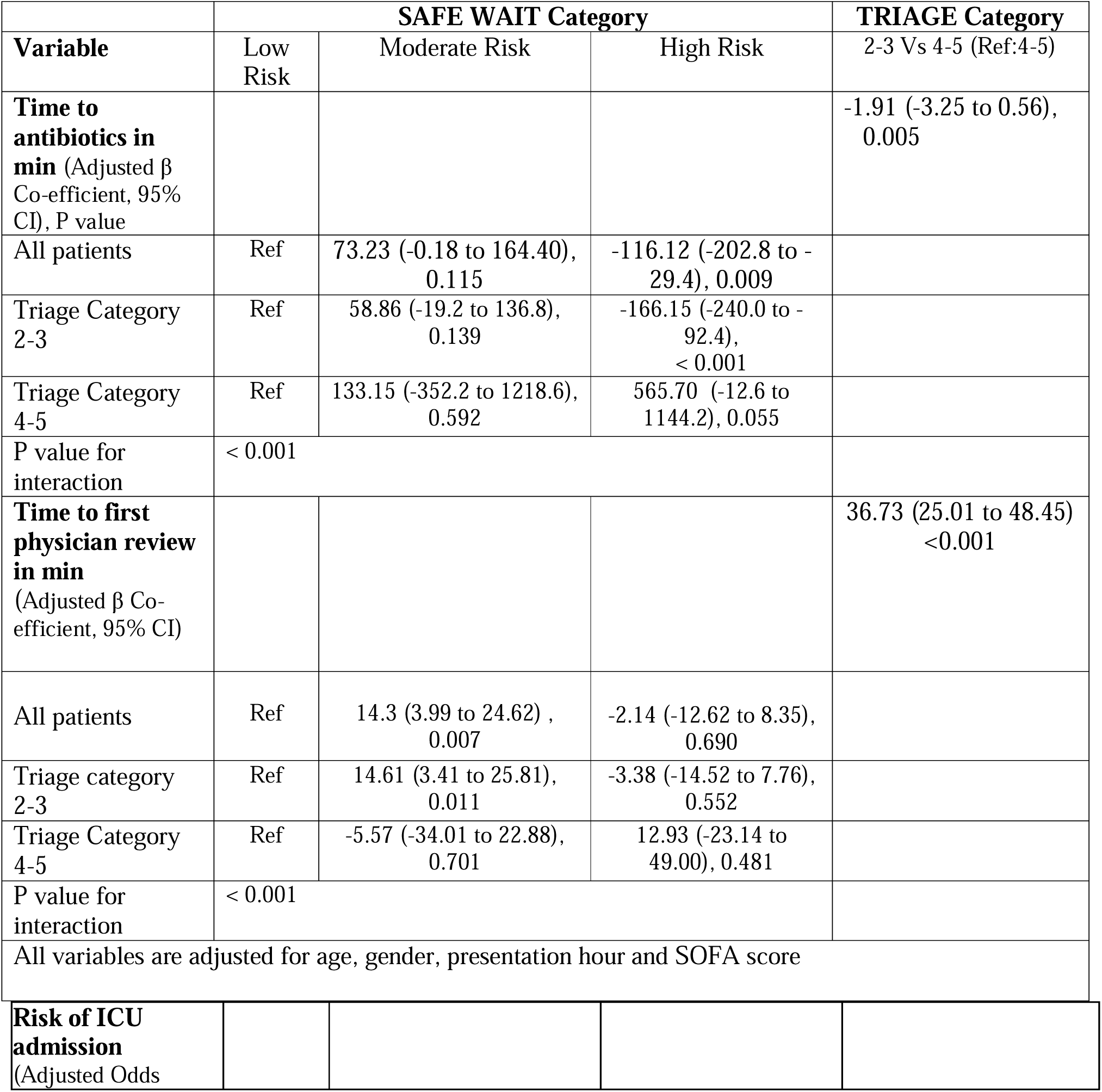
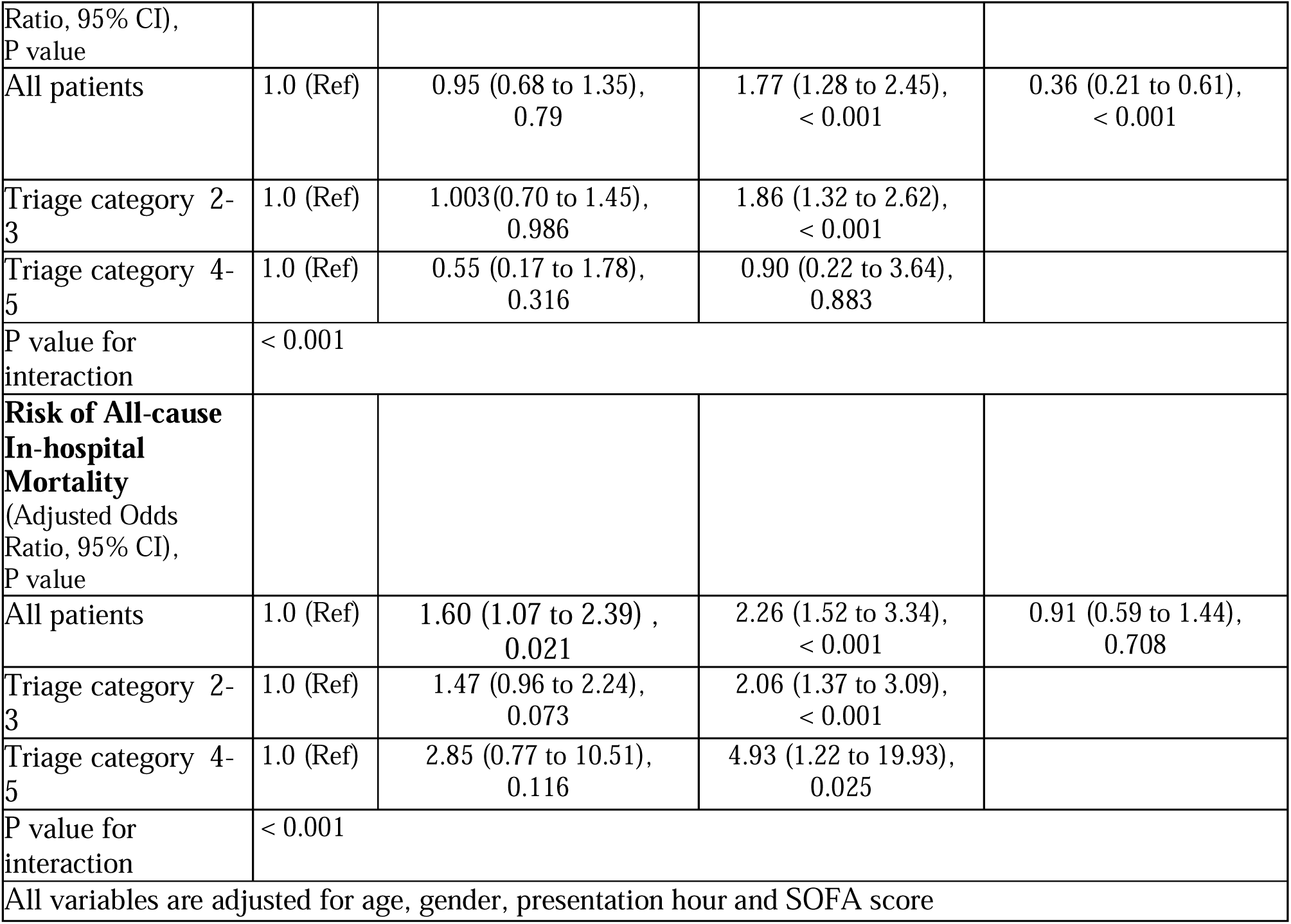
Appendix table. Association between SAFE WAIT category and time to antibiotics, and first physician review, ICU admission and all-cause in-hospital mortality among patients with confirmed sepsis

|  |  | SAFE WAIT Category |  | TRIAGE Category |
| --- | --- | --- | --- | --- |
| Variable | Low Risk | Moderate Risk | High Risk | 2-3 Vs 4-5 (Ref:4-5) |
| Time to antibiotics in min (Adjusted $\beta$ Co-efficient, 95% CI), P value | | | | -1.91 (-3.25 to 0.56), 0.005 |
| All patients | Ref | 73.23 (-0.18 to 164.40), 0.115 | -116.12 (-202.8 to -29.4), 0.009 |  |
| Triage Category 2-3 | Ref | 58.86 (-19.2 to 136.8), 0.139 | -166.15 (-240.0 to -92.4), < 0.001 |  |
| Triage Category 4-5 | Ref | 133.15 (-352.2 to 1218.6), 0.592 | 565.70 (-12.6 to 1144.2), 0.055 |  |
| P value for interaction | < 0.001 |  |  |  |
| Time to first physician review in min (Adjusted $\beta$ Co-efficient, 95% CI) | | | | 36.73 (25.01 to 48.45) <0.001 |
| All patients | Ref | 14.3 (3.99 to 24.62) , 0.007 | -2.14 (-12.62 to 8.35), 0.690 |  |
| Triage category 2-3 | Ref | 14.61 (3.41 to 25.81), 0.011 | -3.38 (-14.52 to 7.76), 0.552 |  |
| Triage Category 4-5 | Ref | -5.57 (-34.01 to 22.88), 0.701 | 12.93 (-23.14 to 49.00), 0.481 |  |
| P value for interaction | < 0.001 |  |  |  |
| All variables are adjusted for age, gender, presentation hour and SOFA score |  |  |  |  |
| Risk of ICU admission (Adjusted Odds |  |  |  |  |
| Ratio, 95% CI),<br>P value |  |  |  |  |
| All patients | 1.0 (Ref) | 0.95 (0.68 to 1.35),<br>0.79 | 1.77 (1.28 to 2.45),<br>< 0.001 | 0.36 (0.21 to 0.61),<br>< 0.001 |
| Triage category 2-3 | 1.0 (Ref) | 1.003(0.70 to 1.45),<br>0.986 | 1.86 (1.32 to 2.62),<br>< 0.001 |  |
| Triage category 4-5 | 1.0 (Ref) | 0.55 (0.17 to 1.78),<br>0.316 | 0.90 (0.22 to 3.64),<br>0.883 |  |
| P value for interaction | < 0.001 |  |  |  |
| **Risk of All-cause In-hospital Mortality**<br>(Adjusted Odds Ratio, 95% CI),<br>P value |  |  |  |  |
| All patients | 1.0 (Ref) | 1.60 (1.07 to 2.39) ,<br>0.021 | 2.26 (1.52 to 3.34),<br>< 0.001 | 0.91 (0.59 to 1.44),<br>0.708 |
| Triage category 2-3 | 1.0 (Ref) | 1.47 (0.96 to 2.24),<br>0.073 | 2.06 (1.37 to 3.09),<br>< 0.001 |  |
| Triage category 4-5 | 1.0 (Ref) | 2.85 (0.77 to 10.51),<br>0.116 | 4.93 (1.22 to 19.93),<br>0.025 |  |
| P value for interaction | < 0.001 |  |  |  |
| All variables are adjusted for age, gender, presentation hour and SOFA score |  |  |  |  |

## Notes

### Competing Interest Statement

The authors have declared no competing interest.

### Funding Statement

Nil funding

### Author Declarations

Ethics approval was obtained from the Western Sydney Local Health District Ethics Committee (Reference numbers 2019/ETH09607 and 2020/ETH03269).

